# NEW EFFECTIVE METHOD OF SPERMATOZOA RECOVERY FROM SURGICALLY-RETRIEVED TESTICULAR AND EPIDIDYMAL SPECIMENS BY DIFFERENTIAL CENTRIFUGATION

**DOI:** 10.1101/2022.11.03.22281832

**Authors:** Zakharova Elena

## Abstract

The newly proposed method of processing cell suspensions for spermatozoa recovery is based on differential centrifugation and allows obtaining male germ cells from biopsy samples and using them for fertilization, especially if they are critically low in number and conventional methods for sperm recovery do no work or are inefficient.

## INTRODUCTION

Of all infertility cases in couples, approximately half is due to male infertility. One of the most severe male factors is azoospermia, a condition when there are no sperm in the man’s semen. If obtaining spermatozoa from the ejaculate is impossible, the next option is to obtain them from the epididymis or testicle, using a number of surgical treatments. In patients with secretory azoospermia, using a surgical microscope, or micro-TESE, is recognized as the most effective method.

At present, it is not the surgical but laboratory stage that ensures the success of the entire treatment. At this stage spermatozoa need to be isolated from the obtained material for further fertilization. Yet there is no universal approach towards single testicular spermatozoa. The efficacy of the laboratory stage depends on laboratory quality and on how much sperm is obtained [1].

Conventional methods of processing testicular and epididymal specimens are based on simple washing (Fig.1). The method is based on settling all cellular elements, including spermatozoa, by centrifugation. After centrifugation, the supernatant fluid is removed and spermatozoa are picked up from the sediment for fertilization by ICSI [2].

Yet in severe cases, for instance, of non-obstructive azoospermia (NOA), isolating single spermatozoa out of testicular tissue is a serious challenge due to their critically low number. Testicular specimens are always contaminated with a large number of non-spermatogenic cells and erythrocytes, often with epithelial spermatogenic cells and cellular debris. Single spermatozoa get stuck and lost in an enormous sediment after centrifugation. Their isolating might be extremely difficult or even impossible (Fig.2A).

We were first to use the method of differential centrifugation for isolating spermatozoa from testicular or epididymal specimens [3]. Differential centrifugation is based on the fact that particles of different size and weight settle at different rates. Larger and heavier particles go first, while smaller and lighter ones remain in the supernatant (Fig. 1). As spermatozoa are smaller in mass than other cells, they can be effectively separated from all other cells in a specimen (Fig. 2B).

**Fig. 1.**
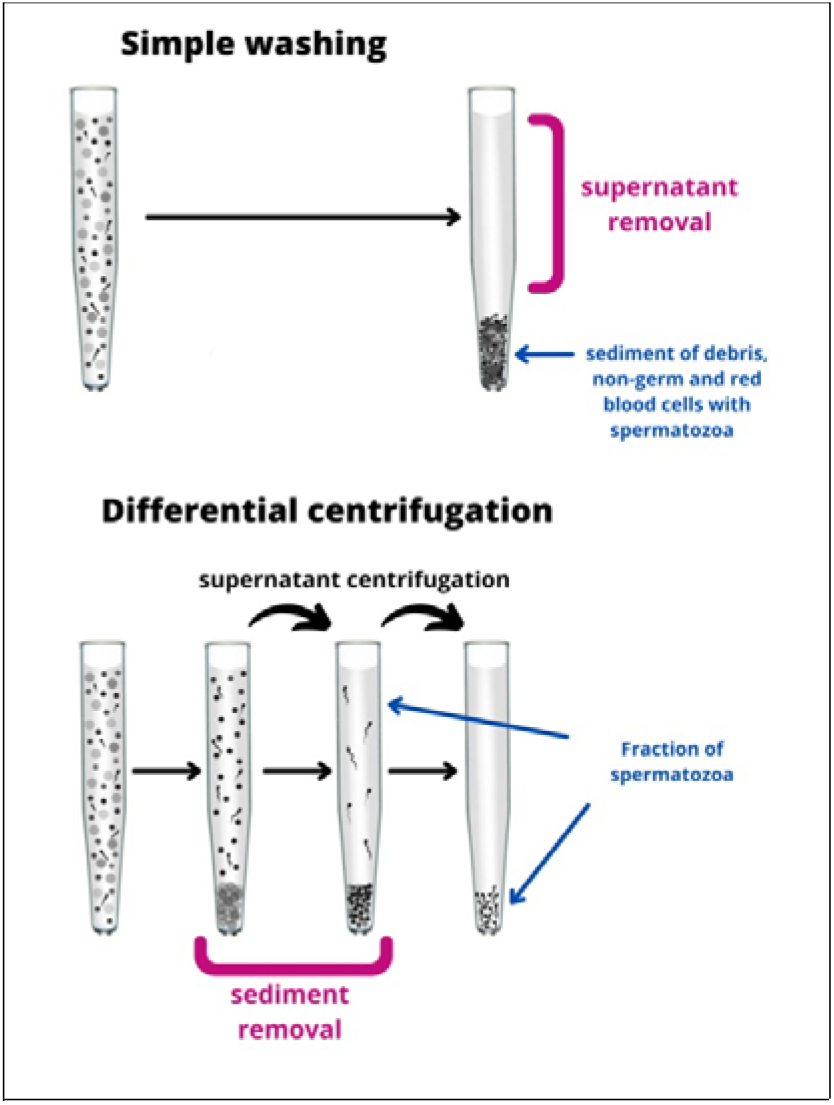
Principles of specimens treatment.

**Fig. 2A.**
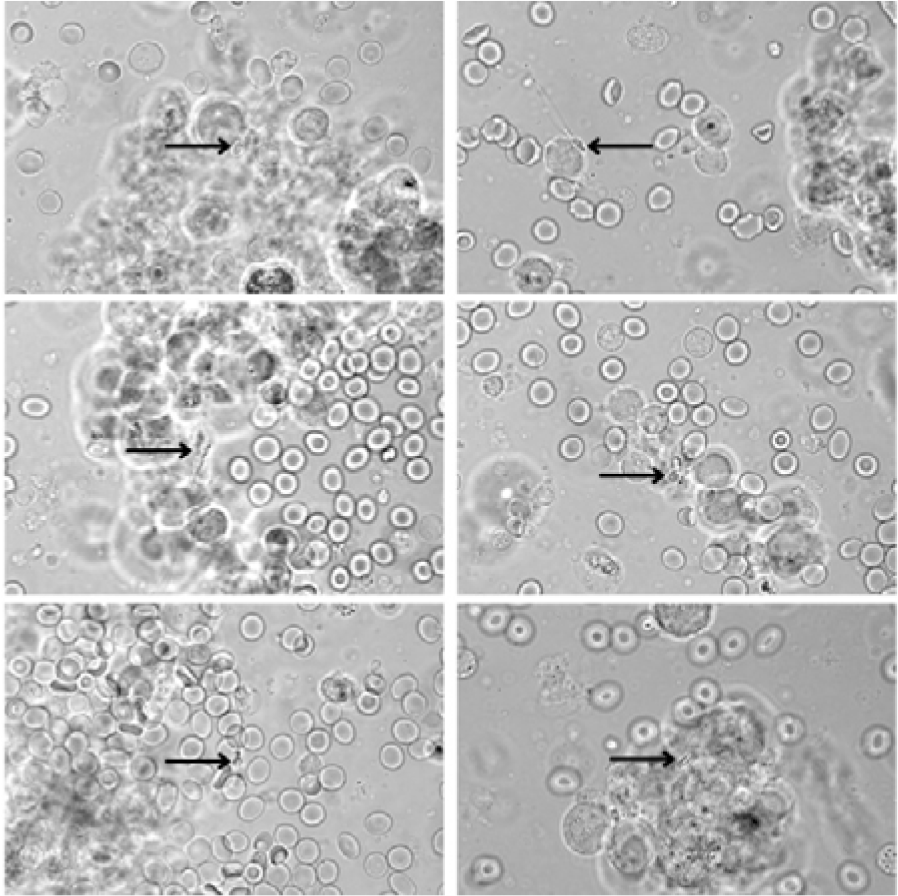
Cell sediment after simple washing, with spermatozoa indicated by arrows.

**Fig.2B.**
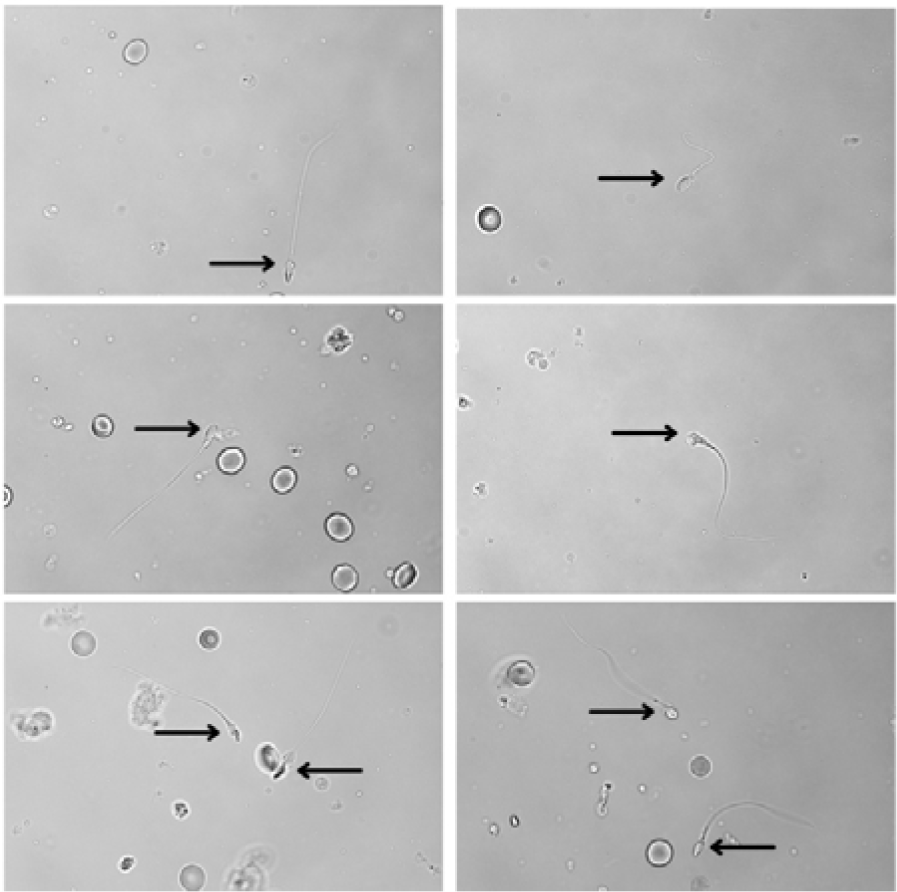
Pure fraction of spermatozoa, indicated by arrows, retrieved after differential centrifugation: the same sample.

## METHODS

A total of 223 testicular open biopsies (micro-TESE) was performed in patients with NOA, and 34 epididymal aspirations/biopsies (MESA/micro-MESE) was performed in patients with obstructive azoospermia (OA).

The biopsy material was first processed mechanically. In all cases, the prepared suspension (or aspirate) is centrifugated for one minute. After centrifugation, the sediment is removed, and the supernatant is re-centrifugated. On average, 8 to 12 successive centrifugation cycles are performed, until there is no visible sediment left. In the last cycle, the supernatant is centrifuged for 10-15 minutes to obtain a concentrated fraction of sperm (Fig. 3). The mode of differential centrifugation is chosen depending on the initial quality of a specimen such as a number of spermatozoa and the degree of contamination with cellular material (on average 250-350 g for 1-2 min, 8-12 times).

**Fig. 3.**
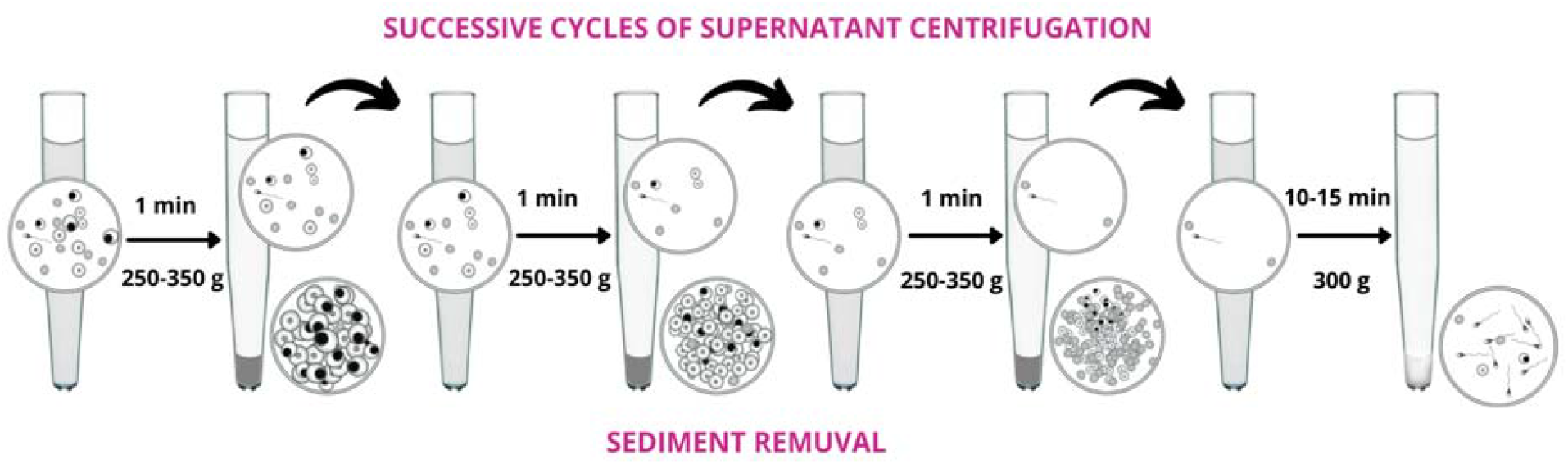
Testicular and epididymal specimens treatment by differential centrifugation.

The recovered spermatozoa were used to fertilize the fresh spouse’s oocytes and/or cryopreserved ones for delayed usage in the IVF/ICSI program. Surplus embryos obtained in IVF programs were cryopreserved for delayed transfer.

## RESULTS

The outcomes of method applying are shown in Table 1.

**Table 1.**
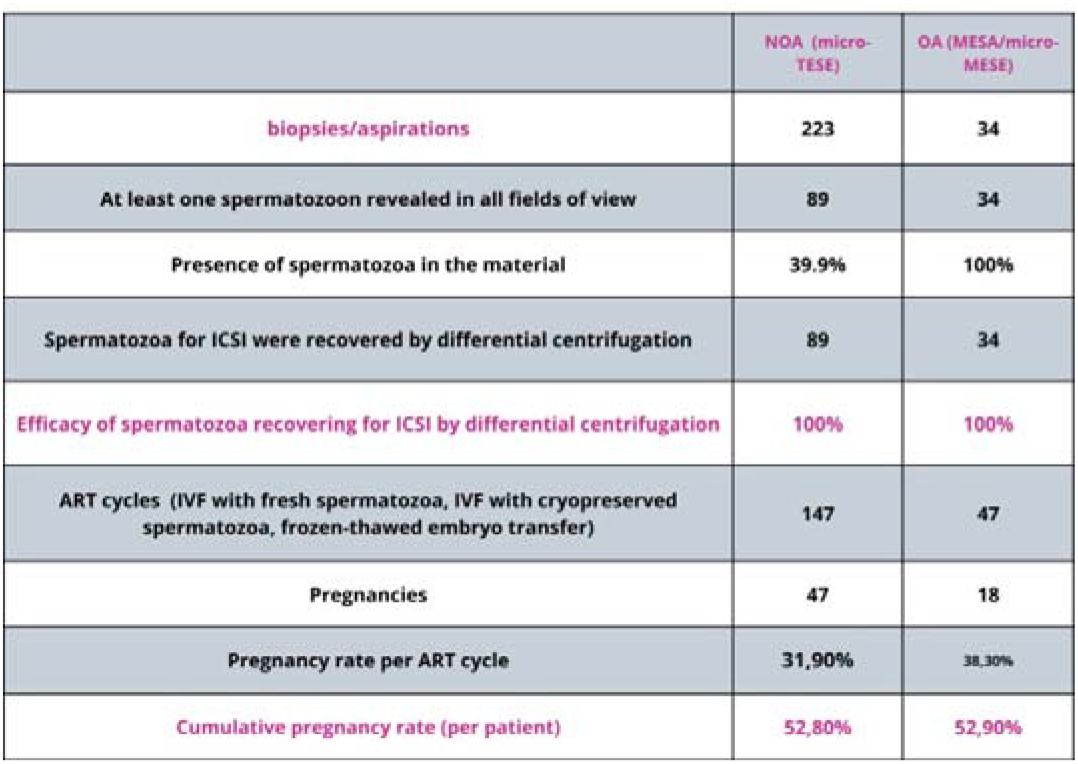
Clinical data.

|  | NOA (micro-TESE) | OA (MESA/micro-MESE) |
| --- | --- | --- |
| biopsies/aspirations | 223 | 34 |
| At least one spermatozoon revealed in all fields of view | 89 | 34 |
| Presence of spermatozoa in the material | 39.9% | 100% |
| Spermatozoa for ICSI were recovered by differential centrifugation | 89 | 34 |
| Efficacy of spermatozoa recovering for ICSI by differential centrifugation | 100% | 100% |
| ART cycles (IVF with fresh spermatozoa, IVF with cryopreserved spermatozoa, frozen-thawed embryo transfer) | 147 | 47 |
| Pregnancies | 47 | 18 |
| Pregnancy rate per ART cycle | 31,90% | 38,30% |
| Cumulative pregnancy rate (per patient) | 52,80% | 52,90% |

In patients with NOA the microscopic examination of cell suspensions, mechanically processed, revealed at least one spermatozoon in all fields of view in 89 patients (39.9%). The further processing of cell suspensions by differential centrifugation was 100% efficient: spermatozoa for ICSI were recovered in all the 89 cases. Pregnancy occurred in 47 out of 89 patients, in 147 ART cycles (52.8% per patient, 31.9% per ART cycle).

In patients with OA, microscopic examination found spermatozoa in the obtained material in all 34 cases (100%). After differential centrifugation, spermatozoa for ICSI and/or cryopreservation were isolated with a 100% efficiency. Pregnancy occurred in 18 out of 34 patients, in 47 ART cycles (52.9% per patient, 38.3% per ART cycle).

## CONCLUSION

The newly suggested method enables isolating even single male germ cells out of aspiration or biopsy material for further fertilization. It is especially useful when their number is critically low and conventional methods might be ineffective.

## Data Availability

All data produced in the present study are available upon reasonable request to the authors

